# Towards Scalable Biomarker Discovery in Posttraumatic Stress Disorder: Triangulating Genomic and Phenotypic Evidence from a Health System Biobank

**DOI:** 10.1101/2025.02.27.25322886

**Authors:** Younga Heather Lee, Yingzhe Zhang, Ana Lucia Espinosa Dice, Josephine H. Li, Justin D. Tubbs, Yen-Chen Anne Feng, Tian Ge, Adam X. Maihofer, Caroline M. Nievergelt, Jordan W. Smoller, Karestan C. Koenen, Andrea L. Roberts, Natalie Slopen

**Affiliations:** Center for Genomic Medicine, Massachusetts General Hospital, Boston, MA; Department of Psychiatry, Harvard Medical School, Boston, MA; Broad Trauma Initiative, Broad Institute of MIT and Harvard, Cambridge, MA; Department of Epidemiology, Havard T. H. Chan School of Public Health, Boston, MA; Diabetes Unit, Department of Medicine, Massachusetts General Hospital, Boston, MA; Programs in Metabolism and Medical and Population Genetics, Broad Institute of Harvard and MIT, Cambridge, MA; Department of Medicine, Harvard Medical School, Boston, MA; Institute of Health Data Analytics and Statistics, College of Public Health, National Taiwan University, Taipei, Taiwan; Center for Precision Psychiatry, Department of Psychiatry, Massachusetts General Hospital, Boston, MA; Department of Psychiatry, University of California San Diego, La Jolla, CA; Veterans Affairs San Diego Healthcare System, Center of Excellence for Stress and Mental Health, San Diego, CA; Veterans Affairs San Diego Healthcare System, Research Service, San Diego, CA; Department of Environmental Health, Havard T. H. Chan School of Public Health, Boston, MA; Department of Social and Behavioral Sciences, Havard T. H. Chan School of Public Health, Boston, MA; Center on the Developing Child, Harvard University, Cambridge, MA

## Abstract

**Importance:** Biomarkers can potentially improve the diagnosis, monitoring, and treatment of posttraumatic stress disorder (PTSD). However, PTSD biomarkers that are scalable and easily integrated into real-world clinical settings have not been identified.

**Objective:** To triangulate phenotypic and genomic evidence from a health system biobank with a goal of identifying scalable and clinically relevant biomarkers for PTSD.

**Design, setting, and participants:** The analysis was conducted between June to November 2024 using genomic samples and laboratory test results recorded in the Mass General Brigham (MGB) Health System. The analysis included 23,743 European ancestry participants from the nested MGB Biobank study.

**Exposures:** The first exposure was polygenic risk score (PRS) for PTSD, calculated using the largest available European ancestry genome-wide association study (GWAS), employing a Bayesian polygenic scoring method. The second exposure was a clinical diagnosis of PTSD, determined by the presence of two or more qualifying PTSD phecodes in the longitudinal electronic health records (EHR).

**Main outcomes and measures:** The primary outcomes were the inverse normal quantile transformed, median lab values of 241 laboratory traits with non-zero *h^2^_SNP_*estimates.

**Results:** Sixteen unique laboratory traits across the cardiometabolic, hematologic, hepatic, and immune systems were implicated in both genomic and phenotypic lab-wide association scans (LabWAS). Two-sample Mendelian randomization analyses provided evidence of potential unidirectional causal effects of PTSD liability on five laboratory traits.

**Conclusion and relevance:** These findings demonstrate the potential of a triangulation approach to uncover scalable and clinically relevant biomarkers for PTSD.

**Key points:** *Question:* Is genetic liability or diagnosis of PTSD associated with clinical laboratory measures?

*Findings:* Among European ancestry participants in the MGB Biobank, we identified 16 unique laboratory traits in cardiometabolic, hematologic, hepatic, and immune systems that were both genetically and phenotypically associated with PTSD, with five markers demonstrating potential unidirectional causal effects of PTSD liability.

*Meaning:* Our findings reveal associations between PTSD and laboratory markers across multiple physiological systems, with evidence suggesting possible unidirectional causal effects of PTSD liability on cardiometabolic, hematologic, and hepatic markers.

## Introduction

Previous epidemiologic investigations of posttraumatic stress disorder (PTSD) have identified several potential biomarkers, primarily focusing on those related to the immune system. For instance, a recent meta-analysis reported elevated levels of inflammatory markers—such as C-reactive protein, interleukin6, and tumor necrosis factor-α—in individuals with PTSD, suggesting a dysregulated immune response as a potential pathophysiological mechanism.^1^ However, by focusing on a narrow range of hypothesis-driven biomarkers, these studies may have overlooked other biological pathways involved in PTSD.

Health system biobanks present a promising alternative for addressing this challenge as they can be linked to large-scale electronic health records (EHR) containing clinical diagnoses, prescriptions, and notably, an extensive array of laboratory test results.^2^ The availability of such rich clinical phenotypes provides an opportunity to capture the complexity of real-world clinical presentations. The integrated data framework enables a comprehensive, hypothesis-free search for biomarkers that are not only scalable but also clinically relevant.

Laboratory-wide association scan (LabWAS) represents an emerging data-driven analytic approach that leverages electronic laboratory measurements to systematically screen a broad spectrum of biomarkers associated with specific target conditions.^3,4^ Initially implemented in cardiovascular disease,^3^ LabWAS has since been adapted to uncover biological and physiological correlates of other complex disease conditions, including major depression,^4^ neuroticism,^5^ and PTSD.^6^ By moving beyond the limitations of targeted, hypothesis-driven approach, LabWAS holds the potential to uncover novel biological pathways involved in PTSD.

In this study, we extend our previous work^3,4^ by integrating data-driven biomarker discovery with comprehensive clinical contextualization and genetic causal investigation. Specifically, we perform two complementary LabWAS analyses using genetic predisposition and clinical diagnosis of PTSD as predictors, leveraging EHR to identify scalable and clinically relevant biomarkers for PTSD. To further elucidate these biomarker-PTSD associations, we evaluate their clinical significance and potential causal mechanisms at the genomic level through two-sample Mendelian randomization (MR) analyses. This integrated approach addresses key limitations of prior studies (e.g., limited clinical relevance and causal insights) and provides a robust framework for discovering novel biomarkers that may ultimately enhance the diagnosis and management of PTSD.^7^

## Methods

### Sample Description

The Mass General Brigham (MGB) Biobank is a hospital-based research program launched in 2010 to support genomic and translational research for human health.^8^ Participants are patients at MGB-affiliated hospitals older than age 18 years at the time of the recruitment who provided informed consent to join the Biobank. Each participant was asked to provide blood samples, which are then linked to their EHR (see Boutin et al., 2022 for further description on enrollment and sample collection).^8^ Given the limited sample size of non-European ancestry participants in the MGB Biobank,^8^ we restricted our analysis to participants of genetically determined European ancestry (see **eMethods** for details of our genotyping, genomic quality control (QC), imputation, and population assignment procedures). Among the European-ancestry participants, our final analytic sample included 23,743 participants with both high-quality genotype and laboratory test results (see **eFigure 1** for the sample flowchart and **eTables 2-10** for sample sizes for each analysis). This research was conducted under a protocol approved by the MGB Institutional Review Board (# 2019P003696).

### Primary Exposure: Bayesian Polygenic Risk Score (PRS) for PTSD (hereafter, PRS_PTSD_)

We used PRS-CS-Auto^9^, a Bayesian polygenic risk scoring method, to generate PRS for European ancestry participants. The weights for PRS were derived from the summary statistics from the most recent genome-wide association studies (GWAS) of PTSD by the Psychiatric Genomics Consortium,^10^ excluding the MGB Biobank samples. We then calculated PRS for each participant by summing all risk-associated variants, weighted by their posterior effect size estimates inferred by PRS-CS-Auto, using PLINK, version 2.0.^11^ Each PRS was subsequently adjusted for sex assigned at birth, age, and the top 10 ancestry principal components to adjust for potential confounding, and standardized to a mean of 0 and a standard deviation of 1.

### Secondary Exposure: PTSD Diagnosis (hereafter, Dx_PTSD_)

We identified PTSD cases by mapping all ICD-9CM and ICD-10CM codes used in MGB-affiliated hospitals to the phecode system^12^ using the PheWAS R package.^13^ We identified two qualifying ICD-9CM codes and five ICD-10CM codes for PTSD (using phecode 300.9; see **eTable 1**). Following our previous approach,^14^ we defined cases as those having at least two qualifying diagnostic codes and the remaining participants as non-cases.

### Outcome: Clinical Laboratory Data

We first extracted clinical laboratory data from the EHR of 2,344,551 MGB patients who self-identified as non-Hispanic White. Subsequently, we preprocessed the laboratory records using the QualityLab pipeline, previously validated in the MGB patient population.^3^ This pipeline implements a series of filtering criteria—including, but not limited to, excluding lab traits with non-numeric values or limited sample sizes (e.g., requiring at least 100 patients, at least 1000 numeric observations), applying unit harmonization (e.g., requiring 70% of the observation measured in the same unit), and applying lab-specific quality control filters (e.g., removing clinically implausible, infinite, or extreme values that are more than four standard deviations from the overall sample mean). Following Dennis et al.’s approach, we extracted the median laboratory value for each patient and applied the inverse normal quantile transformation (INT) to address skewness and non-normality.^3^ Lastly, we calculated SNP-based heritability (*h^2^_SNP_*) for all laboratory tests using restricted maximum likelihood (REML) methods in the genome-wide complex trait analysis (GCTA) software^15^ and identified 241 lab traits with non-zero *h^2^_SNP_* estimates (hereafter, heritable lab traits) for the downstream LabWAS analysis. Further details about the sample size at each stage of preprocessing can be found in **eFigure 1**.

### Laboratory-wide Association Scan (LabWAS)

LabWAS is an innovative, data-driven approach that systematically examines the association between a predictor of interest (e.g., polygenic risk score or diagnostic status) and a wide array of clinical laboratory measurements.^3^ LabWAS involves fitting a series of regression models (equivalent to the number of heritable labs traits resulting from the QualityLab pipeline) that predict the median, INT-transformed values of each heritable lab trait, adjusting for potential confounding factors. In the current study, we estimated associations of PRS_PTSD_ and Dx_PTSD_ with preprocessed lab traits, respectively, adjusting for age at lab measurement, sex assigned at birth (recorded in EHR), and the top 10 genetic PCs (see **eMethods** for details on genomic preprocessing). Subsequently, we identified markers that were statistically significant in both LabWAS analyses after a Bonferroni correction for the number of laboratory tests considered (i.e. 0.05/241).

Recognizing potential heterogeneity in relationships between PTSD (both genetic and phenotypic) and lab traits by patient characteristics, we examined potential effect modification by sex assigned at birth.^16,17^ Additionally, we identified clinically relevant traits that could impact the relationship between PRS_PTSD_ and laboratory markers (see **eFigure 2**), including clinical diagnosis of PTSD, prescription history of selective serotonin reuptake inhibitor (SSRI) recommended to treat PTSD symptoms,^18^ and clinical diagnosis of obesity. Specifically, SSRIs with previously reported metabolic side effects—such as sertraline (RxNorm: 20610), paroxetine (RxNorm: 32937), and fluoxetine (RxNorm: 4493)—may contribute to metabolic dysfunction^19^ by interfering with glucose and lipid metabolism as well as appetite control. Patients with an SSRI prescription history were defined as those with at least one prescription record for these medications. To capture weight gain associated with SSRI use,^20^ we also extracted diagnostic information on obesity (phecode 278) and included it as an additional covariate.

### Two-sample Mendelian Randomization (MR) Analysis

The LabWAS analyses are correlative rather than causal in nature, in part due to the lack of established temporality between PTSD and biomarkers in these data. Thus, following the LabWAS analysis, we performed two-sample MR to assess evidence for possible bidirectional causal relationships between PTSD and 16 unique heritable lab traits that revealed significant associations in both phenotypic and genomic LabWAS analyses (shown in **Table 2**). We used the largest publicly available GWAS summary statistics for PTSD (this time including the MGB samples) and each lab trait (see **eTable 11** for details). We selected genetic instruments for each trait surpassing genome-wide significance (p ≤ 5×10⁻D) and performed clumping to ensure independence (*r²* < 0.001 within a 10,000 kb window) using PLINK v2.0.^11^ We then estimated bidirectional causal effects using multiple complementary methods—including inverse-variance weighted (IVW) regression, MR Egger, maximum likelihood, and weighted median approaches—to explore robustness of results under different model assumptions using the TwoSampleMR R package.^21^

For MR findings to hold causal interpretation, stringent assumptions regarding the instrument(s) and their relationship to exposure and outcome must be met. Specifically, to test the causal null hypothesis (i.e., that exposure does not cause outcome) or to bound the causal effect, the genetic variant(s) used as instrument(s) must: (i) be strongly associated with exposure (i.e., relevance), (ii) have no common cause with the outcome (i.e., independence), and (iii) have effects on the outcome exclusively mediated by the exposure (i.e., exclusion restriction).^22,23^ Only the first assumption may be evaluated empirically; the latter two are unverifiable and must instead be assessed for plausibility through a combination of subject-matter knowledge, falsification tests, and sensitivity analyses.^24–26^ To estimate the magnitude and direction of the effect (i.e., to make meaning of the point estimate), a fourth assumption of either effect homogeneity or monotonicity is required, each with different implications regarding the interpretation of the causal effect estimate.^27^ Given the implications of the latter assumption, rarely mentioned in MR studies,^27–29^ we interpret the magnitude and direction of estimated effects from our two-sample MR with caution. Furthermore, we performed sensitivity analyses for horizontal pleiotropy, directional pleiotropy, and outliers potentially driving pleiotropic biases using the MR-Egger intercept test of deviation from the null, MR-Pleiotropy Residual Sum and Outlier (PRESSO), and leave-1-SNP-out analysis.

## Results

### Sample Characteristics

We first examined participant characteristics in the overall analytic sample and subsamples stratified by the presence of PTSD diagnosis in the EHR. As shown in **Table 1**, the sample was 53.3% female and had a mean age of 61.4 years in 2020. On average, participants with PTSD were significantly younger and more likely to be female (51.7 years of age and 65.7% female) than those without PTSD (61.8 years of age and 52.7% female). PTSD cases were more likely to have public insurance (82.1% vs. 61.4% in non-cases), less likely to be married or live with a partner (31.2% vs. 61.9% in non-cases) and lived in neighborhoods with lower median household income ($70,600) relative to non-cases ($77,100).

**Table 1.** Demographic characteristics of the sample included in the LabWAS analysis.

|  | **Overall** | **PTSD cases** | **Non-cases** | **P-value** |
| --- | --- | --- | --- | --- |
| **Sample size** | 23,743 | 1,045 (4.4%) | 22,698 (95.6%) |  |
| **Mean age in 2020 (SD)** | 61.4 (16.1) | 51.7 (15.1) | 61.8 (16.0) | <0.001 |
| **Sex assigned at birth** |  |  |  | <0.001 |
| Female | 12645 (53.3%) | 687 (65.7%) | 11958 (52.7%) |  |
| Male | 11098 (46.7%) | 358 (34.3%) | 10740 (47.3%) |  |
| **Insurance status** |  |  |  | <0.001 |
| Public payer | 14802 (62.3%) | 858 (82.1%) | 13944 (61.4%) |  |
| Private | 8941 (37.7%) | 187 (17.9%) | 8754 (38.6%) |  |
| **Marital status** |  |  |  | <0.001 |
| Divorced/Separated/Widowed | 3402 (14.3%) | 197 (18.9%) | 3205 (14.1%) |  |
| Married/Partner | 14376 (60.5%) | 326 (31.2%) | 14050 (61.9%) |  |
| Other/Unknown | 255 (1.1%) | 4 (0.4%) | 251 (1.1%) |  |
| Single | 5710 (24.0%) | 518 (49.6%) | 5192 (22.9%) |  |
| **Veteran status** |  |  |  | 0.011 |
| Yes | 2404 (10.1%) | 100 (9.6%) | 2304 (10.2%) |  |
| No or Unknown | 21339 (89.9%) | 945 (90.4%) | 20394 (89.9%) |  |
| **Mean neighborhood median income (SD; in US Dollars)\*** | 76800 (25200) | 70600 (21800) | 77100 (25300) | <0.001 |

\* 277 participants had a missing zipcode and thus had missing information for neighborhood median income.

### Genomic and Phenotypic LabWAS

LabWAS with PRS_PTSD_ revealed statistically significant association with 29 laboratory traits (see **Figure 1a** and **eTable 2**). The strongest associations included higher triglyceride (*ß=*0.08, *p=*4.59×10^-25^), leukocytes (*ß=*0.06, *p=*3.21×10^-21^), glucose (*ß=*0.05, *p=*2.20×10^-15^), alkaline phosphatase (*ß=*0.05, *p=*1.84×10^-13^), and neutrophils (*ß=*0.05, *p=*4.72×10^-13^), erythrocyte distribution width (*ß=*0.04, *p=*4.85×10^-10^), lymphocytes (*ß=*0.04, *p=*6.93×10^-10^), monocytes (*ß=*0.04, *p=*2.22×10^-8^), and hemoglobin A1c (*ß=*0.04, *p=*7.71×10^-8^), as well as lower levels of high-density lipoprotein cholesterol (*ß=*-0.05, *p=*2.04×10^-10^), and potassium (*ß=*-0.04, *p=*3.47×10^-10^).

**Figure 1.**
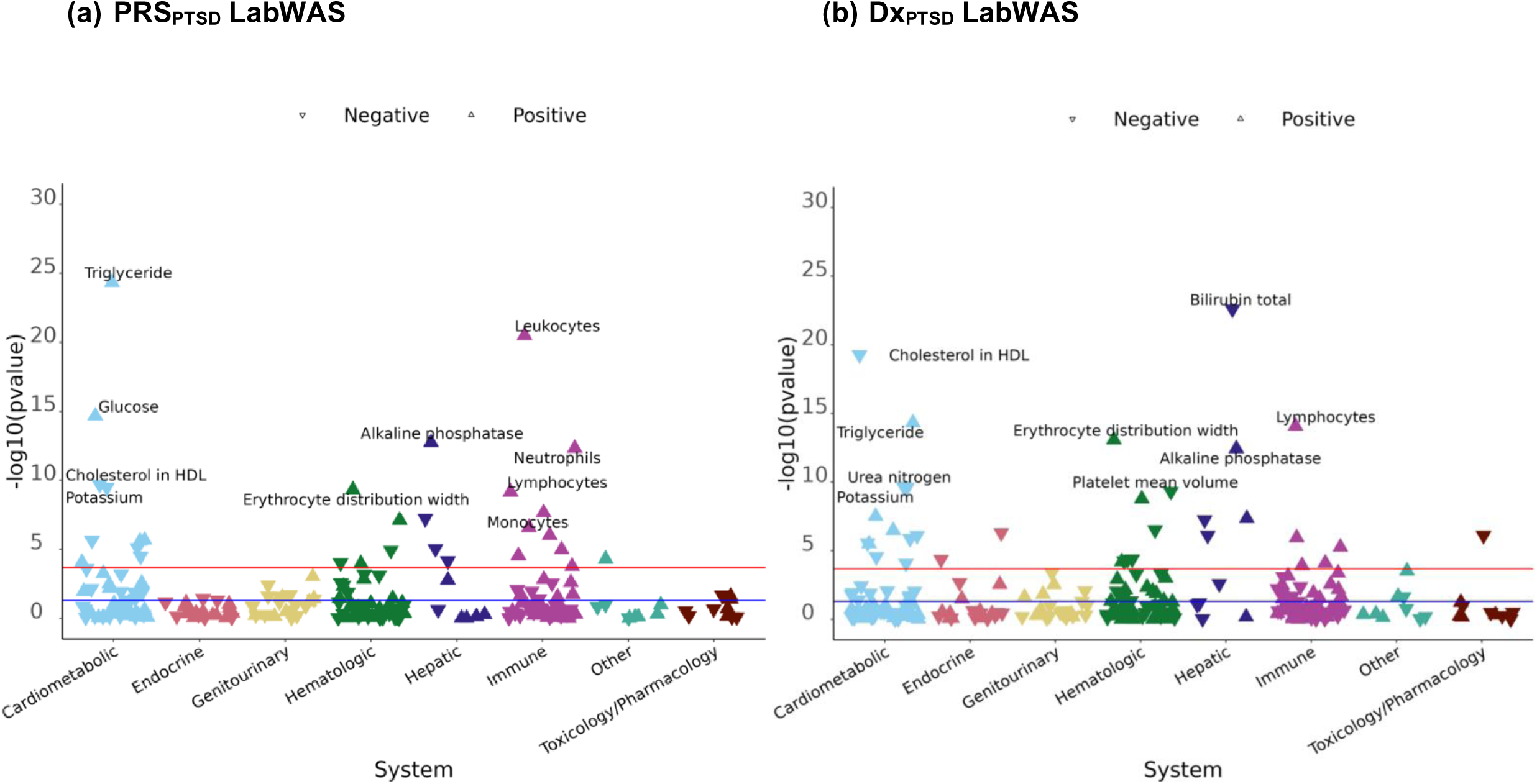
Manhattan plots of lab-wide association scans (LabWAS) using (a) polygenic risk score for posttraumatic stress disorder (PRS_PTSD_) as a predictor (left), and (b) PTSD diagnosis (Dx_PTSD_) as a predictor (right). Lymphocytes* refers to automated lymphocyte count.

LabWAS with Dx_PTSD_ revealed 34 laboratory traits reaching statistical significance (see **Figure 1b** and **eTable 3**), generally with larger effect estimates than seen in the LabWAS with PRS_PTSD_. The strongest associations included higher lymphocytes (*ß=*0.37, *p=*2.72×10^-32^) and triglyceride (*ß=*0.27, *p=*4.72×10^-15^), and lower bilirubin (*ß=*-0.29, *p=*2.52×10^-23^) and high-density lipoprotein cholesterol (*ß=*-0.30, *p=*5.47×10^-20^). Other notable findings included higher erythrocyte distribution width (*ß=*0.22, *p=*8.31×10^-14^), alkaline phosphatase (*ß=*0.23, *p=*3.07×10^-13^), and lower urea nitrogen (*ß=*-0.17, *p=*2.07×10^-10^), potassium (*ß=*-0.19, *p=*2.47×10^-10^), and mean platelet volume (*ß=*-0.37, *p=*4.94×10^-10^).

### Triangulation of Genomic and Phenotypic Evidence

Sixteen unique laboratory markers were statistically significant in both genetic and phenotypic analyses with concordant directions of association (see **Figure 2**, **Table 2**). These overlapping findings spanned multiple biological systems: seven cardiometabolic markers (vitamin D and urea nitrogen had negative associations, while very low-density lipoprotein cholesterol and glucose had positive associations), two hematologic markers (erythrocyte distribution width showing positive association, mean platelet volume showing negative associations), three hepatic markers (albumin, alkaline phosphatase, and bilirubin showing positive associations), and four immune markers (leukocytes, eosinophils, lymphocytes, and monocytes showing positive associations). Notably, three markers (erythrocyte distribution width, high-density lipoprotein cholesterol, triglycerides) showed consistent associations with PRS_PTSD_ and Dx_PTSD_ in sex-stratified analyses, despite reduced statistical power (see **eFigure 2, eTable 4-5** for genomic LabWAS; **eFigure 3, eTable 6-7** for phenotypic LabWAS).

**Figure 2.**
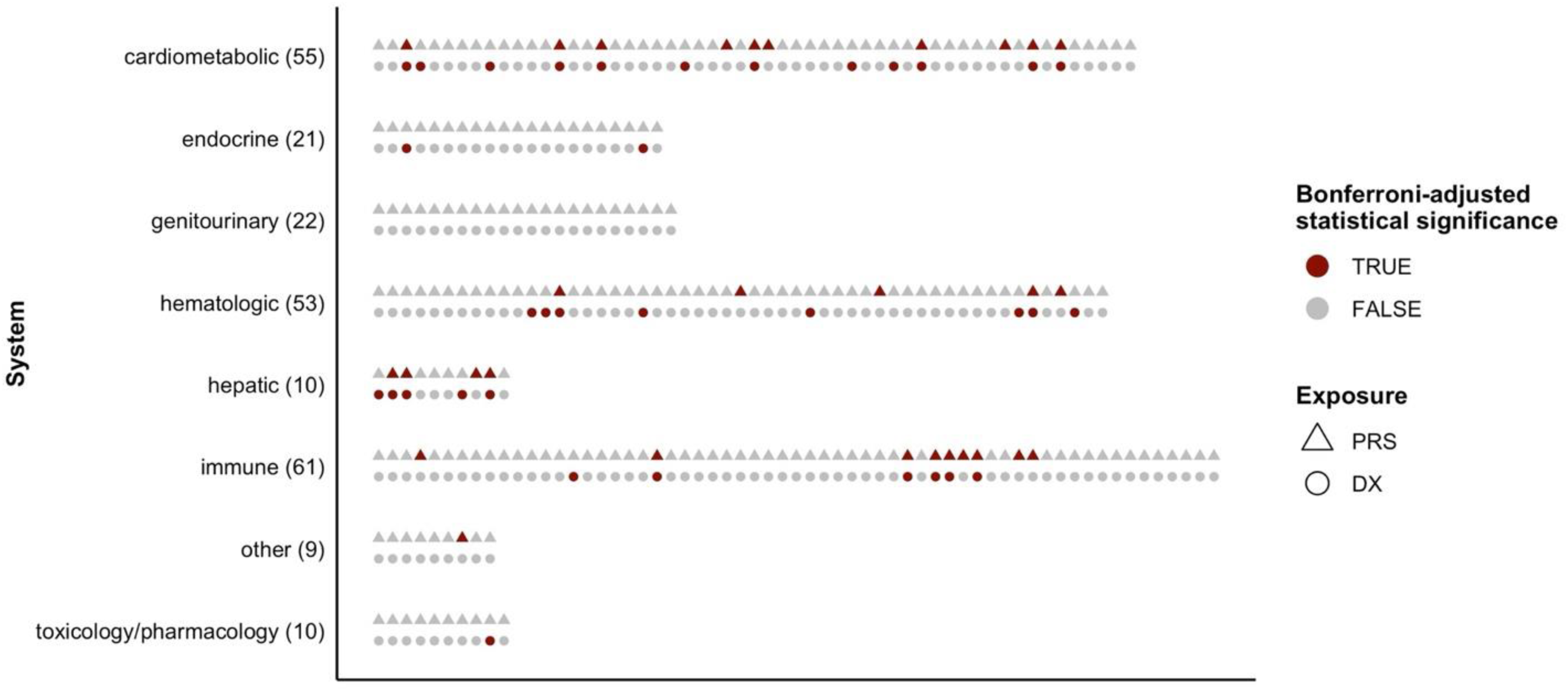
Triangulation of results across PRS_PTSD_ and Dx_PTSD_ LabWAS, grouped by physiological systems. Statistical significance after Bonferroni correction from the genomic LabWAS (shown in triangles) and phenotypic LabWAS (shown in circles), grouped by physiological systems. Physiological systems are sorted alphabetically (top to bottom). Within each system, results are sorted alphabetically (left to right). Number of outcomes per system is listed in parentheses on the y-axis. All sixteen unique markers that are statistically significant in both analyses are concordant in the direction of association. Numeric estimates from the genomic and phenotypic LabWAS can be found in **eTables 2** and **3**, respectively.

**Table 2.** Prior observational and genetic evidence linking PTSD with 16 unique heritable lab traits that were significant in both genomic and phenotypic LabWAS analyses.

| System | Biomarker | Direction of association with PRS<sub>PTSD</sub> and DX<sub>PTSD</sub> | Leading indications and/or reasons for administering the laboratory test<sup>45,46</sup> | Prior <u>observational</u> evidence regarding PTSD and biomarkers | Prior <u>genetically informed</u> evidence regarding PTSD and biomarkers |
| --- | --- | --- | --- | --- | --- |
| Cardio-metabolic | 25-Hydroxyvitamin D3+25 Hydroxyvitamin D2 [Mass/volume] in Serum or Plasma | Negative | Often measured in evaluating risk for osteoporosis and as a screen for deficiency in those who are at high risk due to diet, malabsorption disorder, or kidney or liver disease. | Well-documented associations of PTSD with cardiovascular disease and risk factors, including elevated risk of myocardial infarction and worse lipid profiles (e.g., elevated total cholesterol and triglycerides, lower HDL cholesterol)<sup>33,47–49</sup>; association between PTSD and vitamin D deficiency.<sup>38</sup> | Significant positive genetic correlation ($r_g$) of PTSD with coronary artery disease, hypertension, metabolic syndrome, and heart failure<sup>50–53</sup>; significant association of genetically predicted PTSD with ischemic heart disease, triglycerides, and other circulatory system phenotypes and metabolic laboratory markers<sup>6</sup>; significant bidirectional relationship between PTSD and both coronary artery disease and heart failure that may be discordant in direction<sup>50,51</sup>, as well as significant unidirectional effects of PRS<sub>PTSD</sub> on hypertension and other circulatory and digestive phenotypes<sup>6,51</sup>; possible bidirectional relationship between obesity and PTSD<sup>54</sup>; no causal effect between vitamin D and PTSD based on MR analyses, though two |
|  | Cholesterol in HDL [Mass/volume] in Serum or Plasma | Negative | This test may be part of a routine exam or monitoring for risk and treatment of heart disease (higher HDL cholesterol associated with lower risk of heart disease). |  |  |
|  | Cholesterol in VLDL [Mass/volume] in Serum or Plasma | Positive | Routine part of lipid panel for monitoring risk and treatment of heart disease. |  |  |
|  | Glucose [Mass/volume] in Serum or Plasma | Positive | Routine lab or for risk and treatment monitoring of prediabetes or diabetes. |  |  |
|  | Potassium [Moles/volume] in Serum or Plasma | Negative | Routine part of an electrolyte panel, also used to monitor kidney disease, high blood pressure, heart disease, diabetes, adrenal gland disorders, or medication side effects. |  |  |
|  | Triglyceride [Mass/volume] in Serum or Plasma | Positive | Routine part of lipid panel for monitoring risk and treatment of heart disease. |  |  |
|  | Urea nitrogen [Mass/volume] in Serum or Plasma | Negative | Renal function test, often part of a routine metabolic panel or in evaluation if kidney disease, dehydration, burns, or medication side effects. |  | functional polymorphisms of the vitamin D-binding protein were associated with vitamin D and PTSD.<sup>38,39</sup> |
| Hematologic | Erythrocyte distribution width [Ratio] by Automated count | Positive | Often part of a complete blood count (CBC), measures red blood cell size. May be part of work-up if anemia. High EDW levels may also reflect liver/heart/kidney disease, diabetes, or cancer. | Limited evidence connecting PTSD and these specific blood markers; possible association between PTSD and elevated circulating platelet and red blood cell counts<sup>55</sup>, mean platelet volume<sup>36</sup> and white blood cell count<sup>56</sup> part of proposed biomarker models for PTSD diagnosis/screening. | Genetically regulated expression, colocalization, and local genetic correlation approaches identify associations of genes implicated in PTSD with erythrocyte distribution width, mean platelet volume, and other hematologic markers<sup>35</sup>; significant association of genetically determined PTSD with erythrocyte distribution width and other hematologic markers<sup>6</sup> |
|  | Mean platelet volume [Entitic volume] in Blood by Rees-Ecker | Negative | Often part of a routine CBC, measures platelet size. May be used in evaluation of blood-related conditions, preeclampsia, heart disease, diabetes, cancers, medication side effects, autoimmune diseases, infections, genetic conditions, or alcohol use disorder. |  |  |
| Hepatic | Albumin [Mass/volume] in Serum or Plasma | Negative | Often part of routine liver function panel and may be a marker of nutritional status. Low levels can signal liver, kidney, or digestive disease, malnutrition, infection, or burns. | PTSD associated with viral hepatitis, chronic liver disease, and cirrhosis<sup>57,58</sup>; veterans with PTSD exhibit excess mortality due to viral hepatitis and chronic liver disease<sup>59</sup>; PTSD and substance use share high comorbidity<sup>60</sup>; direct- | Limited large-scale evidence on the shared genetics of PTSD and liver disease; enriched expression of PTSD- and irritable bowel syndrome-shared genes found in liver.<sup>63</sup> |
|  | Alkaline phosphatase [Enzymatic activity/volume] in Serum or Plasma | Positive | Often part of routine liver function panel or used to help diagnose/monitor liver or gall bladder disease, bone disorders, or chronic kidney disease. |  |  |
|  | Bilirubin total [Mass/volume] in Serum or Plasma | Negative | Comprised of both indirect and direct forms of bilirubin, may be used to help diagnose/monitor hepatitis, cirrhosis, other liver diseases, gallbladder disease, bile duct blockage, or hemolytic anemia. | acting antivirals for Hepatitis C treatment associated with improvement in PTSD symptoms<sup>61,62</sup>; alkaline phosphatase part of proposed biomarker model for PTSD screening<sup>56</sup> |  |
| Immune | Eosinophils [# /volume] in Blood | Positive | Part of routine CBC with differential. Also used to evaluate allergic and inflammatory diseases, infections, cancer, blood disorders, autoimmune diseases, or adrenal gland deficiency. Low count may reflect alcohol intoxication or steroid overproduction. | Well-documented associations of PTSD with diseases characterized by immune dysregulation and with elevated proinflammatory markers, including C-reactive protein, interleukins, and white blood cells, but not lymphocytes specifically.<sup>64–66</sup> | Significant $r_g$ of PTSD with C-reactive protein, interleukin-6, white blood cell count, and numerous immune-related phenotypes<sup>67–69</sup>; significant association of genetically determined PTSD with lymphocyte and leukocyte count<sup>6</sup>; significant causal effect of PRS<sub>PTSD</sub> on C-reactive protein and autoimmune thyroid disease<sup>67,68</sup>; evidence regarding causal effect of PRS for immune-related phenotypes/ biomarkers on PTSD (i.e., bidirectional effects) is mixed [unidirectional<sup>67</sup>; bidirectional<sup>6,68</sup>]. |
|  | Leukocytes [# /volume] in Blood by Automated count | Positive | White blood cell count. Also relevant to evaluation immune or inflammatory diseases, infections, cancers, allergic reactions, liver/spleen disease, or tissue or bone marrow damage. |  |  |
|  | Lymphocytes [# /volume] in Blood | Positive | Part of routine CBC with differential. Also used in evaluation of infections and inflammatory diseases, immune diseases, hypothyroidism, blood cancers and diseases, and some inherited conditions. |  |  |
|  | Monocytes [# /volume] in Blood (both manual and automated counts) | Positive | Part of routine CBC with differential. Also used in evaluation of immune or inflammatory diseases, infections, cancers, blood disorders, cardiovascular disease, burn injuries, or aplastic anemia. |  |  |

Several associations from our main analyses were attenuated after adjusting for additional clinical factors (see **eFigure 4**). Three markers associated with PRS_PTSD_ (albumin, platelets, eosinophils) lost significance after adjusting for PTSD diagnosis (comparing **eTable 8** against **eTable 2**), mean corpuscular volume after additionally adjusting for SSRI use (comparing **eTable 9** against **eTable 8**), and three markers (very low-density lipoprotein cholesterol, C-reactive protein, glucose mean value) no longer reached significance after further adjusting for obesity diagnosis (comparing **eTable 10** against **eTable 9**). Notably, the majority of PRS_PTSD_ associations from our primary model remained significant after controlling for PTSD diagnosis, SSRI use, and obesity diagnosis.

### Genomic Causal Inference

Our two-sample MR analyses revealed evidence suggesting unidirectional causal effects. We found no evidence for the causal effects of laboratory traits on PTSD across all MR methods (see **eTable 12**). However, when PTSD liability was modeled as the *exposure*, evidence for statistically significant causal effects emerged across multiple physiological systems (see **eTable 13**). Across all MR methods examined, higher liability for PTSD was causally linked with alterations in hepatic markers, including decreased levels of albumin (IVW *ß* = -3.67×10⁻D, p = 0.0039) and total bilirubin (IVW *ß* = -4.55×10⁻D, p = 5.50×10⁻D). In the cardiometabolic system, PTSD liability exhibited causal relationships with an adverse lipid profile, characterized by decreased high-density lipoprotein cholesterol (IVW *ß* = -4.27×10⁻D, p = 0.00068) and increased very low-density lipoprotein cholesterol (IVW *ß* = 4.65×10⁻D, p = 0.0206). In the hematologic system, higher liability for PTSD showed evidence for causal effects on reduced mean platelet volume (IVW *ß* = -6.59×10⁻D, p = 0.0043), suggesting altered platelet function. While maximum likelihood estimates indicated positive causal effects of PTSD on leukocytes, lymphocytes, and monocytes, these associations were inconsistent across MR methods and warrant cautious interpretation. In summary, our findings suggest that while laboratory traits do not causally impact PTSD, liability for PTSD may induce specific physiological changes— particularly in hepatic, cardiometabolic, and hematological systems—that could represent biological consequences of the disorder.

## Discussion

Our findings identified clinically relevant laboratory markers spanning across cardiometabolic, hematologic, hepatic, and immune systems. These results align with existing research, which has similarly implicated biomarkers—especially immune and metabolic dysregulations—in individuals having genetic predispositions for PTSD^6^ as well as those having PTSD diagnosis.^30,31^ We additionally found potential variations by sex assigned at birth and clinical factors, such as documented PTSD diagnoses and use of psychotropic medications with potential metabolic side effects.^32^ These findings warrant further investigation to better understand the interplay between PTSD, treatments, and subsequent metabolic dysregulations.

Our findings on the association of both PRS_PTSD_ and DX_PTSD_ with cardiometabolic, hematologic, hepatic, and immune markers not only align with but also extend prior evidence linking PTSD to increased risk of cardiometabolic,^33^ hepatic,^34^ and immunological^1^ conditions. Notably, we uncovered novel relationships between PTSD and hematologic markers, particularly the positive relationships between PRS_PTSD_ and Dx_PTSD_ with erythrocyte distribution width, as well as the negative relationships with mean platelet volume. These findings complement recent studies suggesting a shared genetic architecture of PTSD with erythrocyte distribution width^6,35^ and mean platelet volume,^36^ respectively. Furthermore, the observed association between PTSD and vitamin D is noteworthy given emerging research examining the role of vitamin D in shaping the risk of and treatment of internalizing disorders, such as depression and PTSD.^37–39^

The search for biomarkers of PTSD is ongoing, and further research is needed for before these laboratory measures can be used clinically for risk stratification, diagnostics, prognosis, or treatment selection.^40^ Decades of evidence have linked PTSD to widespread physiological dysregulation, including disruptions in stress response system (hypothalamic-pituitary-adrenal axis and sympathetic nervous system), as well as cardiometabolic and immune functions.^41^ More recently, in line with our approach, a growing body of research has leveraged data-driven strategies to develop scalable methods for generating novel and actionable insights, such as multi-omics phenotyping^35^ and machine learning-based diagnostic panel discovery.^36^

### Strengths and Limitations

Our study has several strengths that contribute to the growing body of research on PTSD biomarkers. First, while military cohorts have been instrumental in advancing PTSD genetics research, our use of a health system-based biobank complements this work by providing a balanced sex distribution and representation of civilian populations. This broader demographic coverage enhances the generalizability of our results. Second, this study examines an extensive array of biomarkers, thereby enhancing the potential for discovering novel biomarker targets at scale. Third, our study leverages laboratory tests routinely ordered in clinical care, facilitating future validation studies in real-world settings. Fourth, as shown in **Table 2**, we provide in-depth contextualization of the biomarkers that show strong associations with PRS_PTSD_ and Dx_PTSD_ by describing the leading clinical indications for these laboratory tests and synthesizing prior phenotypic and genetic evidence in relation to PTSD. Finally, by utilizing two-sample MR, we are able to assess evidence for bidirectional causality —albeit under strong, unverifiable assumptions—using genetic instruments from large-scale GWAS.

However, study findings need to be interpreted in light of several limitations regarding the LabWAS analyses. To create usable data from the high-dimensional, longitudinal laboratory records, we implemented the QualityLab preprocessing pipeline that dramatically reduces dimensionality by extracting the median value from each patient’s repeated laboratory measurements. While this approach facilitates downstream analyses, it prevents the ascertainment of temporal sequences of clinical events. While genomic LabWAS may be less susceptible to this limitation compared to phenotypic LabWAS, it can still be biased if the training GWAS for PRS_PTSD_ introduces other sources of bias, such as selection bias and misclassification of PTSD. Furthermore, the ascertainment of trauma and PTSD is particularly challenging in EHR due to unmeasured factors such as social stigma, challenges in accessing care.^42^ In addition, the DSM-5 criteria for PTSD require a confirmed trauma exposure; thus, inaccuracies in measuring trauma exposure can lead to inaccuracies in diagnosing PTSD. Moreover, the fragmented nature of healthcare utilization, especially among patients who navigate between private practices and tertiary hospitals, further complicates an accurate ascertainment of trauma and PTSD.^43^ Lastly, additional limitations regarding our two-sample MR analyses must be considered. Most importantly, the validity of our MR findings relies on strong assumptions that cannot be directly tested. Of key concern is horizontal pleiotropy— where SNPs influence the outcome through pathways independent of the exposure—which would violate the exclusion restriction assumption and invalidate tests of the causal null hypothesis using the IVW estimator.^27^ In addition, sample overlap in the GWASs of PTSD and laboratory markers may bias our estimates; however, this bias is expected to be small given our strong instruments and moderate sample overlap.^44^

### Future Directions

Future studies should replicate our findings across health systems, particularly those serving populations highly impacted by trauma or PTSD. In addition, future investigations are necessary to establish temporal sequence of biomarker measurement, PTSD diagnosis, and relevant clinical factors (e.g., SSRI prescription, metabolic dysfunction) to derive clinical insights most relevant for a specific translational objective (e.g., biological underpinnings of the target condition or the effects of treatment or comorbid conditions). In addition, analytic approaches to incorporate high-dimensional, temporal trajectories of laboratory measurements could further refine our understanding of PTSD biomarkers. Moreover, to further enhance the clinical utility of PTSD biomarkers for diagnostic tools and personalized treatments, future research should consider evaluating how the actual (i.e. untransformed) laboratory values compare to established clinical reference ranges and more rigorously assessing potential causal mechanisms in the observed associations.

## Supporting information

eMethods

eFigure 1

eFigure 2

eFigure 3

eFigure 4

eTables 1-13

## Data Availability

With appropriate Institutional Review Board approval, Mass General Brigham (MGB) investigators and research groups can access genomic data from over 65,000 MGB Biobank participants at no cost, along with their linked clinical data, health information, and survey responses.

## Acknowledgements

This study would not be possible without the contributions of Mass General Brigham (MGB) patients and Biobank participants. We would also like to thank the research coordinators and the MGB Biobank study for their tremendous effort in participant recruitment and sample collection. Lastly, we would like to acknowledge the MGB Research Patient Data Registry (RPDR) team for their work maintaining the enterprise research patient data warehouse.

