## Supplementary material for "Towards Scalable Biomarker Discovery in Posttraumatic Stress Disorder: Triangulating Genomic and Phenotypic Evidence from a Health System Biobank": eMethods


**eMethods.** Genomic preprocessing procedure at the Mass General Brigham (MGB) Biobank.

**eFigure 1.** Sample selection flow chart.

**eFigure 2.** Comparison of genomic LabWAS results between female and male participants.

**eFigure 3.** Comparison of phenotypic LabWAS results between female and male participants.

**eFigure 4.** Genomic LabWAS results, adjusted for relevant clinical factors, including PTSD diagnosis, medication use, and obesity.

**eTable 1.** ICD codes included in the case definition for posttraumatic stress disorder.

**eTable 2.** Results from genomic LabWAS reaching Bonferroni statistical significance.

**eTable 3.** Results from phenotypic LabWAS reaching Bonferroni statistical significance.

**eTable 4.** Female sex-stratified results from genomic LabWAS reaching Bonferroni statistical significance.

**eTable 5.** Male sex-stratified results from genomic LabWAS reaching Bonferroni statistical significance.

**eTable 6.** Female sex-stratified results from phenotypic LabWAS reaching Bonferroni statistical significance.

**eTable 7.** Male sex-stratified results from phenotypic LabWAS reaching Bonferroni statistical significance.

**eTable 8.** Results from genomic LabWAS, further adjusted for $Dx_{PTSD}$, reaching Bonferroni statistical significance.

**eTable 9.** Results from genomic LabWAS, further adjusted for $Dx_{PTSD}$ and SSRIs, reaching Bonferroni statistical significance.

**eTable 10.** Results from genomic LabWAS, further adjusted for $Dx_{PTSD}$, SSRIs, and $Dx_{Obesity}$, reaching Bonferroni statistical significance.

**eTable 11.** GWAS summary statistics used in the two-sample mendelian randomization (MR) analysis.

**eTable 12.** Results from two-sample MR, modeling PTSD as an outcome.

**eTable 13.** Results from two-sample MR, modeling PTSD as an exposure.

**eMethods.** Genomic preprocessing procedure at the Mass General Brigham (MGB) Biobank.

The MGB Biobank samples (N = 36,424) were genotyped on Multi-Ethnic Global Array kits from Illumina (Illumina Inc., San Diego, USA) and released in eight batches. We performed batch-specific genotype data QC to remove single nucleotide polymorphisms (SNPs) with genotype missing rate >0.05, samples with genotype missing rate >0.02, and SNPs with differential missing rate >0.01 between any two batches, after which different batches were merged for subsequent QC steps.

As MGB Biobank included individuals from diverse populations, we inferred the genetic ancestry of biobank participants using 1000 Genomes samples (1KG) as the population reference panel. Specifically, we computed principal components (PCs) for biobank samples and 1KG samples combined and trained a random forest classifier to assign a "super population" label for biobank samples with a prediction probability ≥0.9 using the first 6 PCs of the 1KG samples as the training data. This resulted in 25,677 individuals whose ancestry was classified as European (EUR), 1,607 as African (AFR), 1,840 as Admixed American (AMR), 504 as East Asian (EAS), and 297 as South Asian (SAS) ancestry. Within each ancestry, we excluded samples with mismatched reported and genetic sex, outliers of the absolute value of heterozygosity (>5 standard deviations from the mean), and one sample from each pair of related individuals (identity-by-descent (IBD) >0.2); SNPs that showed significant batch associations at $P < 1 \times 10{-}4$, had a missing rate > 0.02 or Hardy–Weinberg equilibrium (HWE) test $P < 1 \times 10{-}10$ were also discarded.

Next, we used the Michigan Imputation Server (Minimac4) to impute genotype dosages for biobank samples, with the Haplotype Reference Consortium (HRC) as the reference panel for EUR ancestry. Lastly, we removed markers with imputation quality INFO score <0.8, minor allele frequency (MAF) <0.01, a significant deviation from HWE with $P < 1 \times 10{-}10$, and missing rate >0.02. The dataset uses genome build 37 (hg19). Further information about genotyping, QC, imputation, and population assignment procedures for the MGB Biobank is available on the following GitHub repository (https://github.com/Annefeng/PBK-QC-pipeline).
