## Supplementary figures and images for "Towards Scalable Biomarker Discovery in Posttraumatic Stress Disorder: Triangulating Genomic and Phenotypic Evidence from a Health System Biobank"

### eFigure 1

**eFigure 1.** Sample selection flowchart.

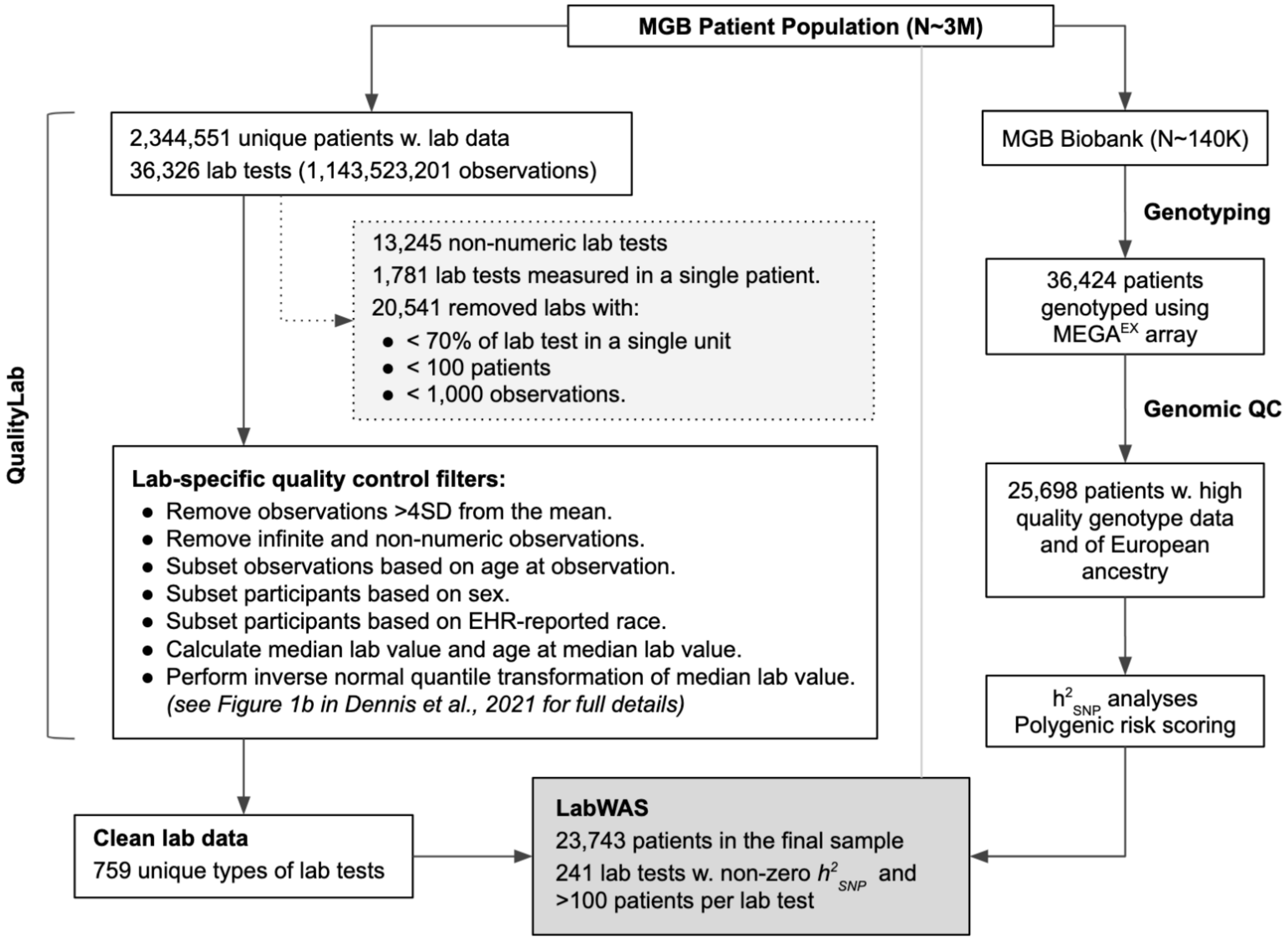
