## Supplementary material for "Towards Scalable Biomarker Discovery in Posttraumatic Stress Disorder: Triangulating Genomic and Phenotypic Evidence from a Health System Biobank": eFigure 3

**eFigure 3.** Comparison of phenotypic LabWAS results between female and male participants.

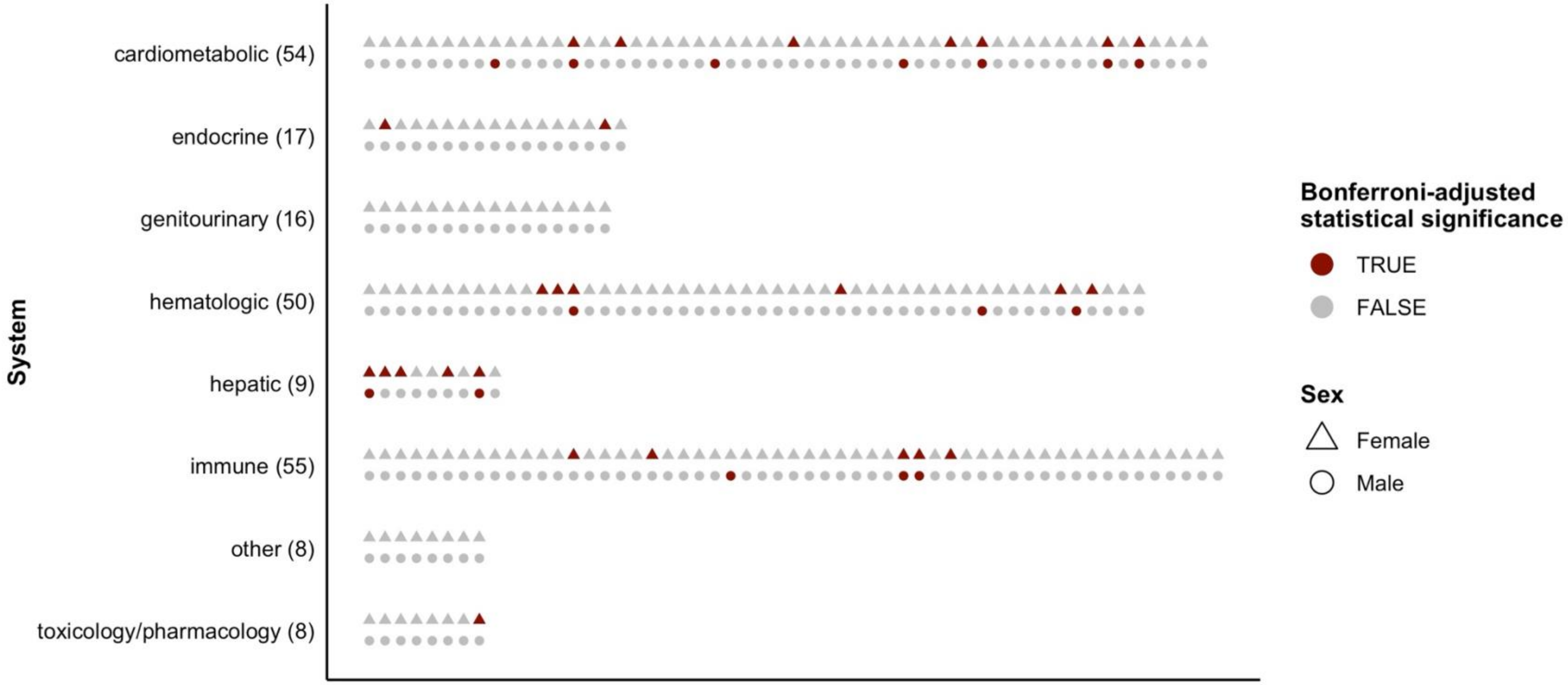


**Caption:** Statistical significance after Bonferroni correction from the $Dx_{PTSD}$ LabWAS among females (triangles) and males (circles), grouped by physiological systems. Systems are sorted alphabetically (top to bottom). Within each system, results are sorted alphabetically (left to right). Number of outcomes per system is listed in parentheses on the y-axis. All outcomes that are statistically significant in both analyses are concordant in the direction of association. Numeric estimates from the female-specific LabWAS and male-specific LabWAS can be found in **eTables 6** and **7**, respectively.
