## Supplementary material for "Towards Scalable Biomarker Discovery in Posttraumatic Stress Disorder: Triangulating Genomic and Phenotypic Evidence from a Health System Biobank": eFigure 4

**eFigure 4.** Genomic LabWAS results, adjusted for relevant clinical factors, including PTSD diagnosis, medication use, and obesity.

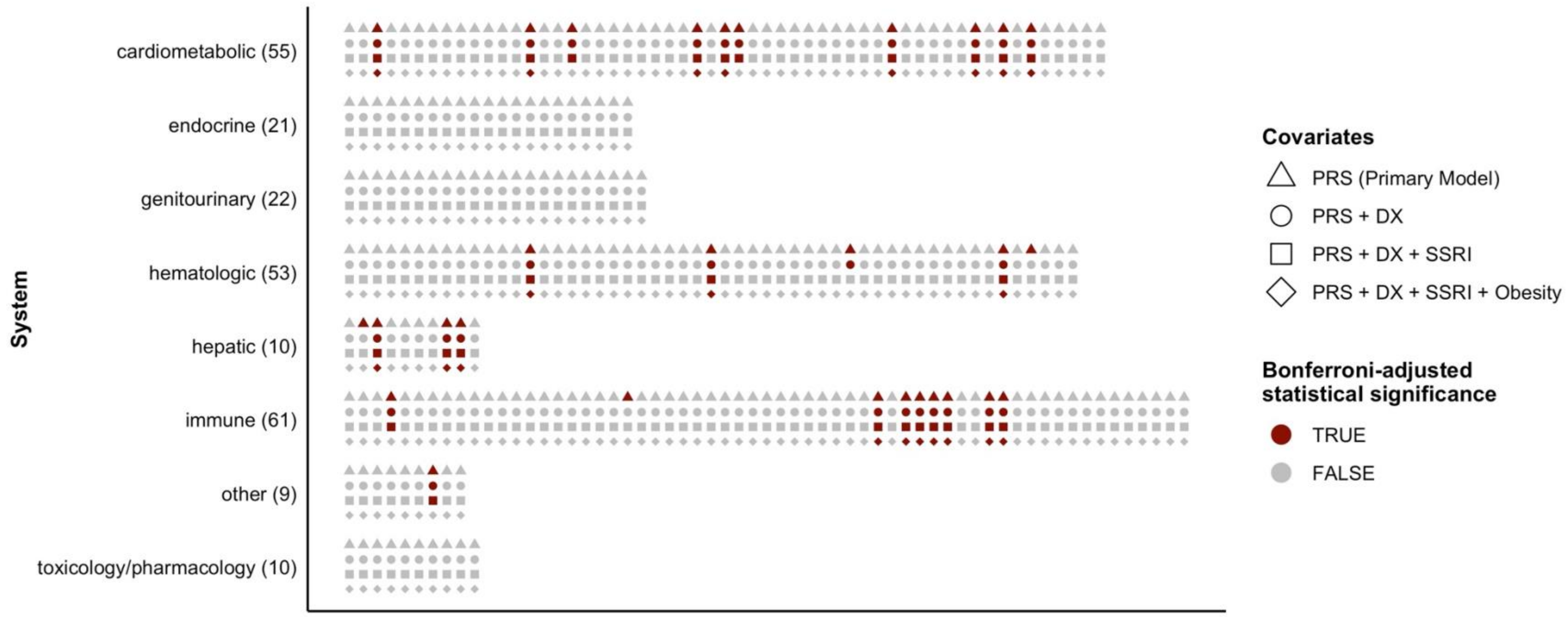


**Caption:** Statistical significance after Bonferroni correction from the $PRS_{PTSD}$ LabWAS as presented in **Figure 2** (triangles) and with additional adjustment for $Dx_{PTSD}$, SSRI, and $Dx_{Obesity}$ (circles, squares, and diamonds, respectively), grouped by physiological systems. Systems are sorted alphabetically (top to bottom). Within each system, results are sorted alphabetically (left to right). Number of outcomes per system is listed in parentheses on the y-axis. All outcomes that are statistically significant in the primary analysis are concordant in the direction of association across these sensitivity analyses. Numeric estimates from each analysis can be found in **eTables 2, 8, 9,** and **10**, respectively.
